# Geographic and temporal trends in etiology-specific diarrhea burden among children in low-resource settings

**DOI:** 10.64898/2026.04.01.26349890

**Authors:** Maria Garcia Quesada, James A. Platts-Mills, Patricia B. Pavlinac, Helen Powell, Karen L. Kotloff, Elizabeth T. Rogawski McQuade

## Abstract

**Background:** Several large multisite studies have been conducted to describe etiology-specific burden of diarrhea among children in low-resource settings. Here, we combined data across studies to describe geographic and temporal trends in incidence and attributable fractions (AFs) of etiology-specific moderate-to-severe diarrhea (MSD), and to evaluate etiology-specific case fatality ratios (CFRs).

**Methods:** We harmonized case definitions and analytic methods across the Global Enteric Multicenter Study (GEMS), Malnutrition and Enteric Disease (MAL-ED), Vaccine Impact on Diarrhea in Africa (VIDA), AntiBiotics for Children with severe Diarrhea (ABCD), and Enterics for Global Health (EFGH) studies. Cases were 6–35-month-olds with acute MSD. Incidence estimates for GEMS, VIDA, and EFGH were adjusted for enrollment, healthcare seeking, and diagnostic testing. AFs were calculated as the proportion of MSD cases attributed to each etiology, and CFRs were estimated within 14 and 90 days of an MSD episode.

**Findings:** Pre-rotavirus vaccine introduction, rotavirus had the highest incidence and was the leading etiology among 6–11-month-olds, accounting for approximately 22–28% of MSD; the proportion of diarrhea due to rotavirus declined following vaccine introduction, with average AF 10–11% in Africa and Asia. *Shigella* incidence was highest among 12-23-month-olds and was the dominant etiology among 12-23 and 24-35-month-olds, causing approximately one-third to one-half of MSD. Overall, 90-day mortality declined substantially over time, from 2.21% in GEMS to 0.30% in EFGH. Bacterial (2.52%) and protozoal pathogens (3.55%) had higher average CFRs than viral pathogens (1.42%).

**Conclusion:** Harmonized analysis of five multisite studies reveals consistent evidence that rotavirus and *Shigella* are the dominant causes of MSD in children under three years in low-resource settings, with burden shifting toward *Shigella* following rotavirus vaccine introduction.

## 1 Background

Morbidity and mortality due to diarrheal disease have substantially declined in the past few decades. In 2021, about 1.17 million deaths globally were attributed to diarrhea, which is a 60% decline since 1990, with most improvements being among children <5 years of age.^1^ These reductions are a result of a combination of etiology agnostic interventions (e.g., improvements in water, sanitation, and hygiene [WASH]; oral rehydration solution [ORS]) and etiology-specific interventions (e.g., immunization, antibiotic access).^2^ Planning for and prioritizing future interventions requires an understanding of how etiology-specific diarrhea has changed over time and geography. However, because diarrhea can be caused by a great variety of pathogens including bacteria, viruses, and protozoa, and the proportion of diarrhea cases that access care is relatively low, etiology-specific disease burden is difficult to characterize. Several large multisite studies of diarrhea among children have been conducted in sub-Saharan Africa, Asia, and Latin America to address this gap, but due to logistical limitations, these studies typically only enrolled for 2-3 years, and they included a limited number of sites. Here, we combined data across studies to elucidate whether etiology-specific burden differs across geography and how it has changed over time, and to evaluate etiology-specific mortality, which is a rare outcome and therefore difficult to assess in any individual study.

First, the Global Enteric Multicenter Study (GEMS) was a population-based, age-stratified prospective case-control study that estimated etiology-specific acute moderate-to-severe diarrhea (MSD) burden among children aged 0–59 months seeking care between 2007 and 2011.^3^ Second, the Etiology, Risk Factors, and Interactions of Enteric Infections and Malnutrition and the Consequences for Child Health and Development (MAL-ED) longitudinal birth cohort study followed children from birth to two years of age between 2009 and 2014.^4^ MAL-ED included both active diarrhea surveillance and monthly testing of non-diarrheal stool samples. Third, the Vaccine Impact on Diarrhea in Africa (VIDA) followed the same study design as GEMS and was conducted between 2015 and 2018.^5^ Fourth, the AntiBiotics for Children with severe Diarrhea (ABCD) study was a randomized clinical trial to estimate the impact of azithromycin treatment on acute non-bloody, watery diarrhea among children ages 2-23 months between 2017 and 2019. Fifth, the Enterics for Global Health (EFGH) *Shigella* surveillance study was a hybrid surveillance study that aimed to describe etiology-specific incidence of *Shigella*, and other pathogens, among children 6-35 months between 2022 and 2024.^6^

Each of these five studies made major contributions to our understanding of etiology-specific diarrheal disease epidemiology among children in low-resource settings. However, to best address their individual objectives, they each used varying case definitions, study designs, and statistical methods such that their published results are not directly comparable. In this study, we attempted to harmonize case definitions and analytic methodology across studies in order to describe trends in incidence and attributable fractions (AF) of etiology-specific MSD over time and geography and to estimate average etiology-specific case fatality ratios (CFR).

## 2 Methods

### 2.1 Harmonization of data

The design and statistical methods of GEMS, MAL-ED, VIDA, ABCD, and EFGH have been described elsewhere.^3–12^ To best understand how trends have changed over time, we aimed to harmonize the case definitions and methods from older studies to match those in EFGH – the most recent study – wherever possible. However, because GEMS and VIDA used a more restrictive case definition than EFGH, we used the MSD case definition from those studies: three or more loose stools within 24 hours, fewer than seven days since the start of diarrhea, and at least one indicator of dehydration (i.e., sunken eyes, loss of skin turgor, or intravenous hydration), dysentery, or hospital admission. MAL-ED captured all community cases of diarrhea using active surveillance and EFGH captured all diarrhea cases that sought care, so we subset cases from those two studies to those that met the GEMS/VIDA MSD case definition. Incidence and AF analyses were stratified by age groups included in EFGH: 6-11 months, 12-23 months, and 24-35 months. For some analyses, countries represented across studies were grouped into regions including Africa (Kenya, Mali, Mozambique, The Gambia, Malawi, Tanzania, South Africa), Asia (Bangladesh, India, Nepal, Pakistan) and Latin America (Brazil, Peru).

We focused on MSD due to the following etiologies: adenovirus (40 and 41), astrovirus, *Campylobacter jejuni* or *Campylobacter coli, Cryptosporidium*, heat-stable toxin producing enterotoxigenic *E. coli* (ST ETEC), norovirus GII, rotavirus, sapovirus, *Shigella*, and typical enteropathogenic E. coli (typical EPEC).^10^ Each study attributed diarrhea etiology differently based on pathogen quantities detected by quantitative PCR (qPCR). Here, we attributed etiologies using qPCR cycle threshold (Ct) cutoffs for whole stool samples as developed in EFGH, which were derived for the primary age range analyzed here (6-35 months);^13^ since EFGH did not involve controls, these cut-offs were derived from GEMS and MAL-ED participants. In EFGH only, cases had a mix of whole stool and/or rectal swab samples that were used for etiologic attribution; for the latter, Ct cutoffs were adjusted to account for different sensitivity. MSD cases attributed to multiple etiologies were counted towards each etiology-specific incidence, AF, and CFR estimate.

GEMS, VIDA, and EFGH were included in all analyses, but ABCD and MAL-ED were only included in a subset of analyses due to data limitations. Since dysentery cases were excluded in ABCD, and these are expected to have a different distribution of etiologies compared to watery diarrhea cases, MSD cases from this study were excluded from incidence and AF analyses. ABCD MSD cases included in CFR analyses were restricted to those that were randomized to placebo since azithromycin treatment likely differentially impacted etiology-specific mortality rates. MAL-ED was included in incidence and AF analyses but did not have results for the 24–35-month age group since participants were only followed through 24 months of age. MAL-ED also could not be included in CFR analyses since qPCR was not performed for children that did not complete follow-up, which includes children who died during the study (n=20).

### 2.2 Incidence

#### 2.2.1 MAL-ED

To estimate incidence of etiology-specific MSD in MAL-ED, a longitudinal birth cohort study, we calculated the total number MSD cases attributed to each etiology during age-specific time periods (6-11 months and 12-23 months). We fit Poisson regression models with country site and age group as predictors and an offset for follow-up time. Incidence rates per 100 child-years and 95% confidence intervals were obtained from model predictions.

#### 2.2.2 GEMS, VIDA, and EFGH

To estimate total incidence of etiology-specific MSD cases in the community in GEMS and VIDA, we replicated the EFGH analytic adjustments applied to enrolled MSD cases as closely as possible. These methods have been described in detail elsewhere.^14,15^ Briefly, MSD cases enrolled in facilities were adjusted for enrollment, healthcare seeking, and diagnostic testing.

To adjust for eligible MSD cases that sought care at a study facility but were not enrolled, we applied two weights. The first upweighted for those who were not enrolled because the site- and age-specific enrollment cap had been met for a given time period (e.g., 2 weeks in GEMS and VIDA). The second upweighted for those who were not enrolled for any other reason (e.g., declined to consent, study staff not available, etc.), and this second weight was stratified by site, age, and watery vs bloody diarrhea.

To adjust for MSD cases that did not seek care at a study facility, we also applied two weights estimated from the healthcare utilization surveys (HUS) administered among the population living in the facility catchment area. The first was a site-specific weight to account for those who sought care at a different facility. The second was an individual-level weight to account for those who did not seek care at any facility. This was estimated using a propensity score model that used disease severity and sociodemographic characteristics as predictors for care seeking. The specification of each variable included varied slightly by study based on the specific questions included in the HUS, but included descriptors of site, age, sex, number of stools, blood in stool, vomiting, fever, and wealth quintile. Of note, the GEMS model did not include wealth quintile because the questions used to construct this indicator were not asked in the abbreviated HUS survey that was used to estimate care seeking among MSD cases.^16^ The models also included two-way interactions selected with LASSO, and individual-level weights were truncated at the 95^th^ percentile as described elsewhere.^14^

To adjust for pathogen testing rates, we applied site- and pathogen-specific weights for enrolled cases that were tested but did not have a valid result for a given pathogen. In GEMS only, only a random subset of cases within site- and age-specific strata underwent qPCR testing, so we also applied an additional weight to account for those that were not tested.^8^

Population denominators were estimated via census in GEMS and VIDA and required no adjustment. We assumed a uniform age distribution to estimate denominators for finer age groups (e.g., 6-11 months from 0–11-month census, and 24-35 months from 24–59-month census). In EFGH, the enumerated population was adjusted as previously described.^17^

Lastly, adjusted incidence was estimated by dividing the adjusted number of MSD cases by the population denominator, and percentile 95% confidence intervals around the adjusted estimate were estimated via bootstrap as done in EFGH.^14^

### 2.3 Attributable Fractions and Case Fatality Ratios

AFs were calculated as the proportion of MSD cases etiologically attributed to each pathogen, stratified by study, country site, and age group. MSD cases from GEMS, VIDA, and EFGH underwent the same adjustments as for incidence calculations described above. Given similar AF distributions across sites and studies within each rotavirus vaccine introduction status group (pre- and post-introduction), we also calculated weighted average AFs stratified by region, rotavirus vaccine introduction status, and age group. We weighed each study-site (i.e., each country site within each study) AF equally to avoid overrepresentation of country sites or studies with larger sample sizes.

CFRs were estimated as the proportion of MSD cases attributed to each pathogen and pathogen class (i.e., viruses, bacteria, and protozoa) that died within 14-or 90-days. CFRs for bacteria also included *Salmonella, V. cholerae*, and *Aeromonas*, and CFRs for protozoa also include *Isospora, E. histolytica*, and *Cyclospora*. Due to small numbers, we estimated CFRs by collapsing cases across all studies and sites, and included all MSD cases 0-59 months, acknowledging ABCD and EFGH did not enroll this full age range. As a sensitivity analysis, we estimated CFRs restricted to the age range in EFGH (6-35 months) and excluding ABCD given even finer age range and differing eligibility criteria of cases. We also estimated 90-day CFRs for MSD cases who were hospitalized at enrollment since these estimates may be used for mortality estimates that rely on the etiology distribution of hospitalized cases.

Confidence intervals were calculated using stratified bootstrap resampling with 1000 iterations. For each bootstrap iteration, children were resampled with replacement within each study-site stratum to preserve the original distribution of sample sizes across sites. Percentile-based 95% confidence intervals were constructed from the bootstrap distribution.

## 3 Results

### 3.1 Study Characteristics and Sample Size

This analysis combined data from five multi-site diarrhea studies conducted between 2007 and 2024 across sites in three continents by sampling enrollees across varying time frames and populations who met harmonized case definitions. GEMS (2007-2011) enrolled 9,439 MSD cases (58.9% with qPCR results for all top 10 pathogens) from seven countries across Africa and Asia. MAL-ED (2009-2014) captured 989 MSD cases (97.4% with qPCR results) among its birth cohort from eight countries in Africa, Asia, and Latin America. VIDA (2015-2018) enrolled 4,480 MSD cases (95.3% with qPCR results) in three African countries. ABCD (2017-2019) enrolled 1,806 MSD cases (97.2% with qPCR results) into the placebo arm in seven countries in Africa and Asia. The most recent study, EFGH (2022-2024), enrolled 3,098 MSD cases (93.4% with qPCR results) in seven countries across Africa, Asia, and Latin America. For countries where multiple studies were conducted, the population catchment area was not always consistent across studies and therefore may not be comparable (Table 1).

**Table 1.**
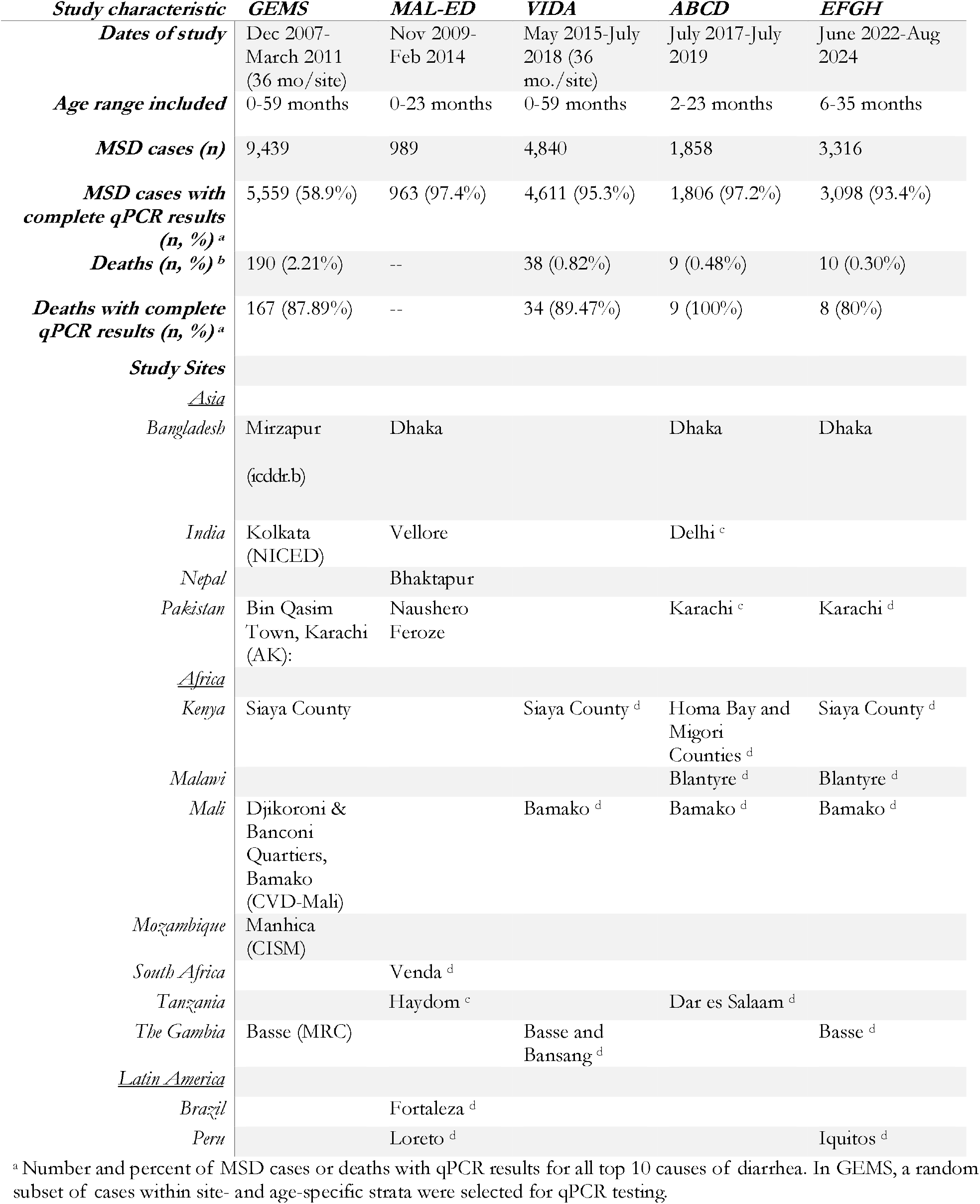
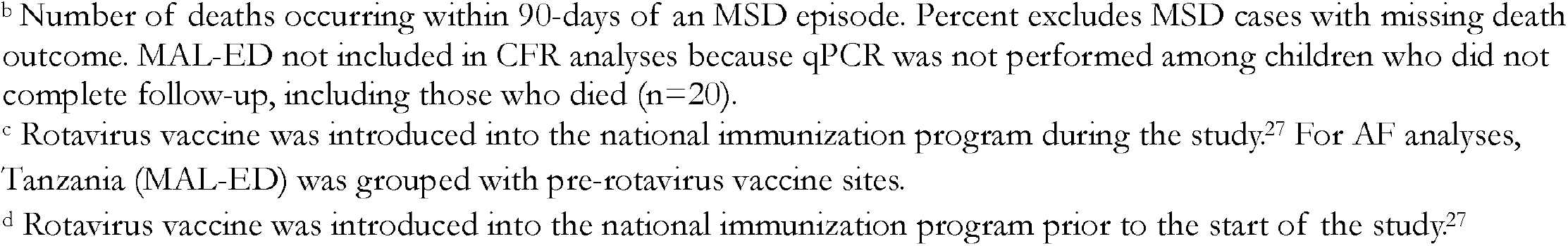
Dates, age ranges, MSD cases, deaths, and sites included in each study.

### 3.2 Incidence

The incidence of MSD varied by country site and study but was consistently highest among children 6-11 months old and then decreased with age (Supplementary Table 1). Within sites, there was no consistent pattern in MSD incidence over time (i.e., across studies; Supplementary Table 1). Rotavirus MSD incidence, which was also highest among children 6-11 months, appeared to decline following rotavirus vaccine introduction. In Mali (Bamako), rotavirus vaccine was introduced between GEMS and VIDA; rotavirus MSD incidence among children 6-11 months declined from 22.11 cases per 100 person years (95% CI: 13.45, 37.75) in GEMS, to 7.54 (4.58, 18.04) in VIDA and 3.71 (1.99, 8.13) in EFGH (Figure 1). This trend was consistent in The Gambia and Pakistan, but not Kenya (Figure 1). Other countries did not have both pre- and post-rotavirus vaccine introduction data. Contrary to rotavirus, *Shigella* MSD incidence was generally highest among children 12-23 months compared to 6-11-or 24-35-month-olds, and it appeared to decline over time in some country sites but not all (Figure 1). Incidence of MSD due to all etiologies by study, country site, and age group are available at https://mariagq.shinyapps.io/diar-trends-dashboard/.

**Figure 1.**
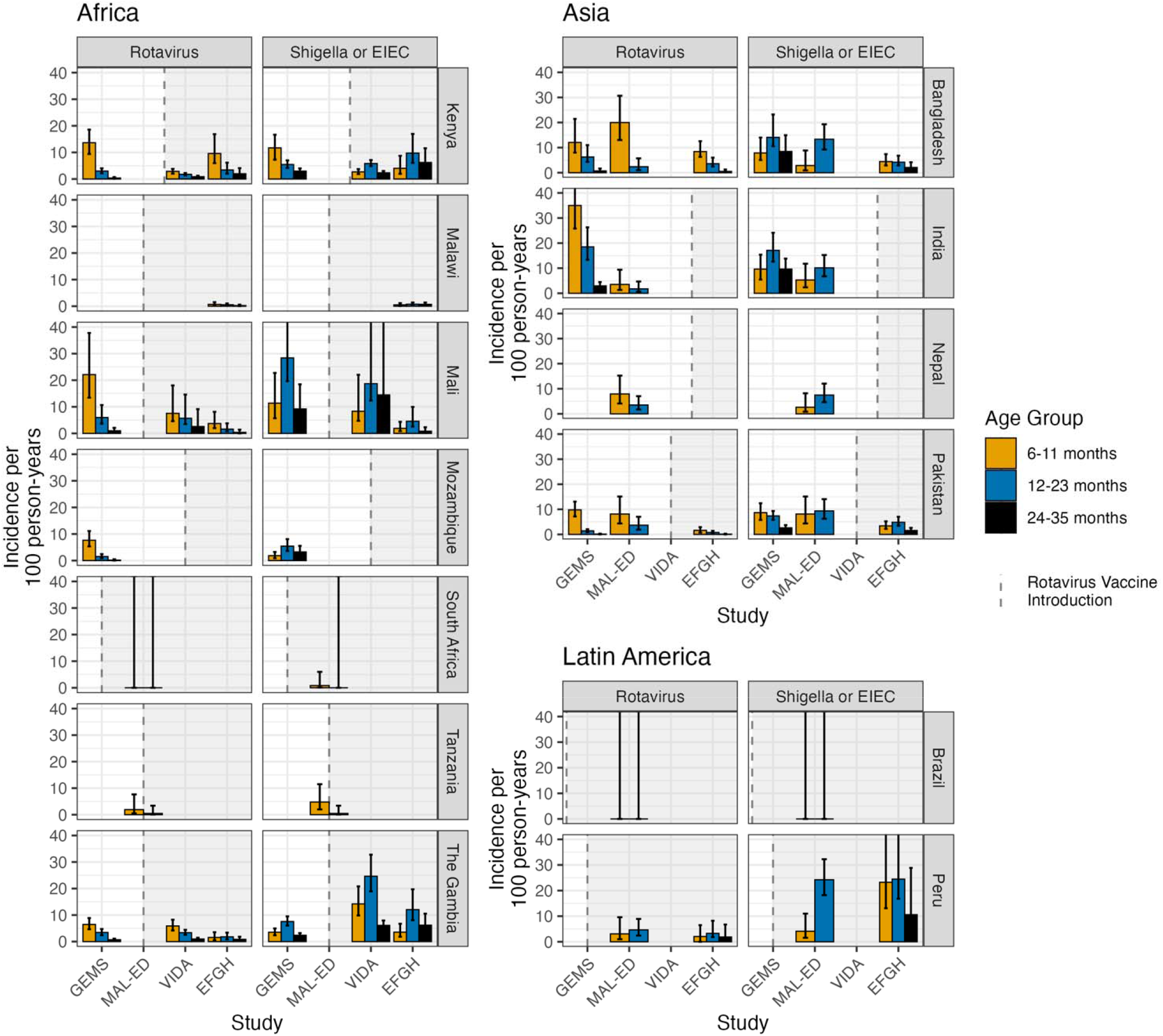
Age-specific adjusted incidence of rotavirus and *Shigella* MSD by country across studies.

### 3.3 Attributable Fractions

The leading etiology of MSD shifted after rotavirus vaccine introduction among 6–11-month-olds. Prior to rotavirus vaccine introduction, rotavirus was the leading etiology among 6–11-month-olds on average, followed by *Shigella* and ST ETEC (Figure 2). Following rotavirus vaccine introduction, the average proportion of MSD attributed to rotavirus among 6–11-month-olds went from 22.40% (95% CI: 17.97, 27.17) to 9.74% (8.07, 12.11) in Africa, and from 28.35% (24.26, 32.80) to 11.05% (5.50, 17.89) in Asia (Figure 2; Supplementary Table 2). In Latin America, where only post-rotavirus vaccine data was available, 2.85% (0.74, 5.61) of MSD was attributed to rotavirus among 6–11-month-olds (Figure 2; Supplementary Table 2). This shift was also observed in individual studies and sites; rotavirus was the leading etiology among 6–11-month-olds in 9/13 study-sites prior to vaccine introduction, and the leading etiology varied after vaccine introduction (*Shigella* in 4/11, *Campylobacter jejuni or C. coli* in 2/11, *Cryptosporidium* in 2/11, rotavirus in 2/11, and ST ETEC in 1/11; Supplementary Table 3).

**Figure 2.**
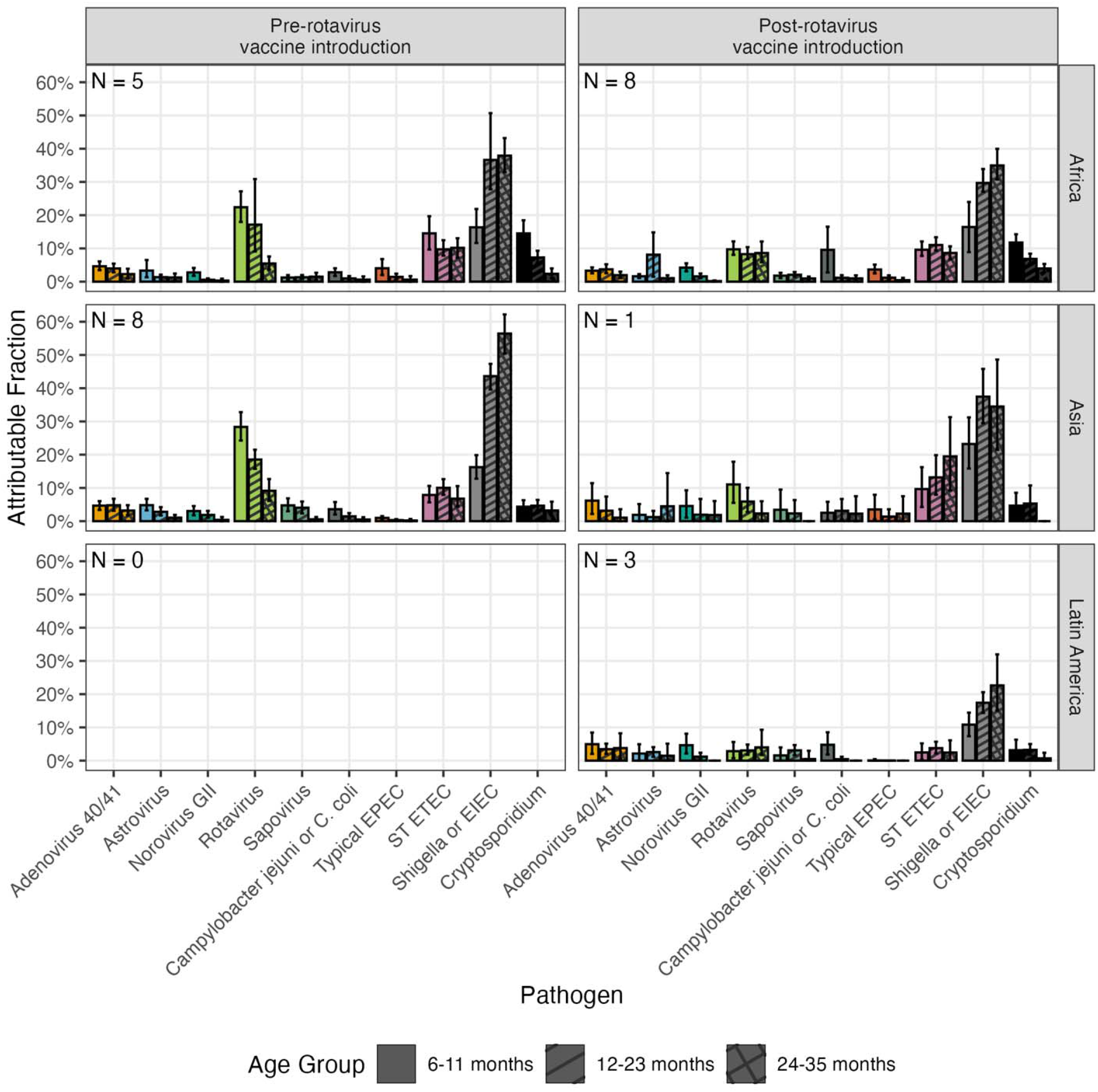
Distribution of AFs averaged across studies and sites pre- and post-rotavirus vaccine introduction into the national immunization program. N in each plot corresponds to the number of study-sites (i.e., country sites in each study) included in that strata. Studies include GEMS, MAL-ED, VIDA, and EFGH.

*Shigella* was the leading etiology of MSD among 12–23- and 24–35-month-olds, both pre- and post-rotavirus vaccine introduction. Prior to rotavirus vaccine introduction, about a third of MSD was attributed to *Shigella* in Africa (36.65% [27.79, 50.71] among 12–23-month-olds, 37.92% [32.96, 43.21] among 24–35-month-olds) and close to half in Asia (43.62% [39.62, 47.32] among 12–23-month-olds, 56.45% [50.41, 62.20] among 24–35-month-olds), on average (Figure 2; Supplementary Table 2). Similarly, in individual studies and country sites, *Shigella* was the leading etiology in 11/13 study-sites among 12–23-month-olds and in all 8/8 study-sites among 24-35-month-olds (Supplementary Table 3). Post-rotavirus vaccine introduction, the average proportion of MSD due to *Shigella* among 12–23 and 24–35-month-olds was slightly lower but remained close to a third in both Africa and Asia (Figure 2). Data from Latin America was sparse and only available in post-rotavirus vaccine settings, but *Shigella* was the leading etiology in those study-sites among 12–23- and 24–35-month-olds, as well as in almost all African and Asian study-sites (9/11 study-sites for 12-23-month-olds and 9/9 study-sites for 24-35-month-olds; Supplementary Table 3).

ST-ETEC was also an important pathogen in Africa and Asia, causing 6.77-19.49% of MSD on average across age group and rotavirus vaccine status strata, and its prevalence did not follow a clear age pattern (Figure 2; Supplementary Table 2). *Cryptosporidium* was also an important cause of MSD in Africa, particularly among younger children, causing 14.52% (11.18, 18.47) and 11.78% (9.81, 14.27) of MSD among 6–11-month-olds pre- and post-rotavirus vaccine introduction, respectively, on average (Figure 2; Supplementary Table 2). *Cryptosporidium* was less common in Asia (0.00-4.67%), and neither *Cryptosporidium* (0.68%-3.24) nor ST-ETEC (2.43-3.77%) were very common in Latin America across age groups (Figure 2; Supplementary Table 2). AFs for all etiologies by study, country site, and age group are available at https://mariagq.shinyapps.io/diar-trends-dashboard/.

### 3.4 Case Fatality Ratios

Overall 90-day mortality following MSD has declined over time, from 2.21% in GEMS, 0.82% in VIDA, 0.48% in ABCD, and 0.30% in EFGH (Table 1). Etiology-specific CFRs, calculated across all studies and age groups, revealed significant variation by etiology. Typical EPEC was associated with the highest 90-day CFR at 8.75% (95% CI: 5.06, 12.61), followed by *Cryptosporidium* at 3.76% (2.44, 5.27; Figure 3). Restricting the age range from 0-59 to 6-35 months, and excluding ABCD cases, did not meaningfully change the results (Supplementary Figure 1). As a group, viral pathogens had a lower 90-day CFR (1.42% [0.97,2.01]) than bacterial (2.52% [2.02, 3.06]) or protozoal (3.55% [2.34, 4.96]) pathogens (Figure 3). The 14-day CFRs followed similar patterns but were lower than 90-day estimates (Figure 3). Etiology-specific 90-day CFRs for hospitalized cases were imprecise due to few deaths; pathogen-class 90-day CFRs for hospitalized cases were higher but followed a similar trend as overall CFRs (3.78% [2.05, 6.10] for viruses, 10.06% [7.64, 12.72] for bacteria, and 12.95% [7.38, 18.20] for protozoa; Supplementary Table 4).

**Figure 3.**
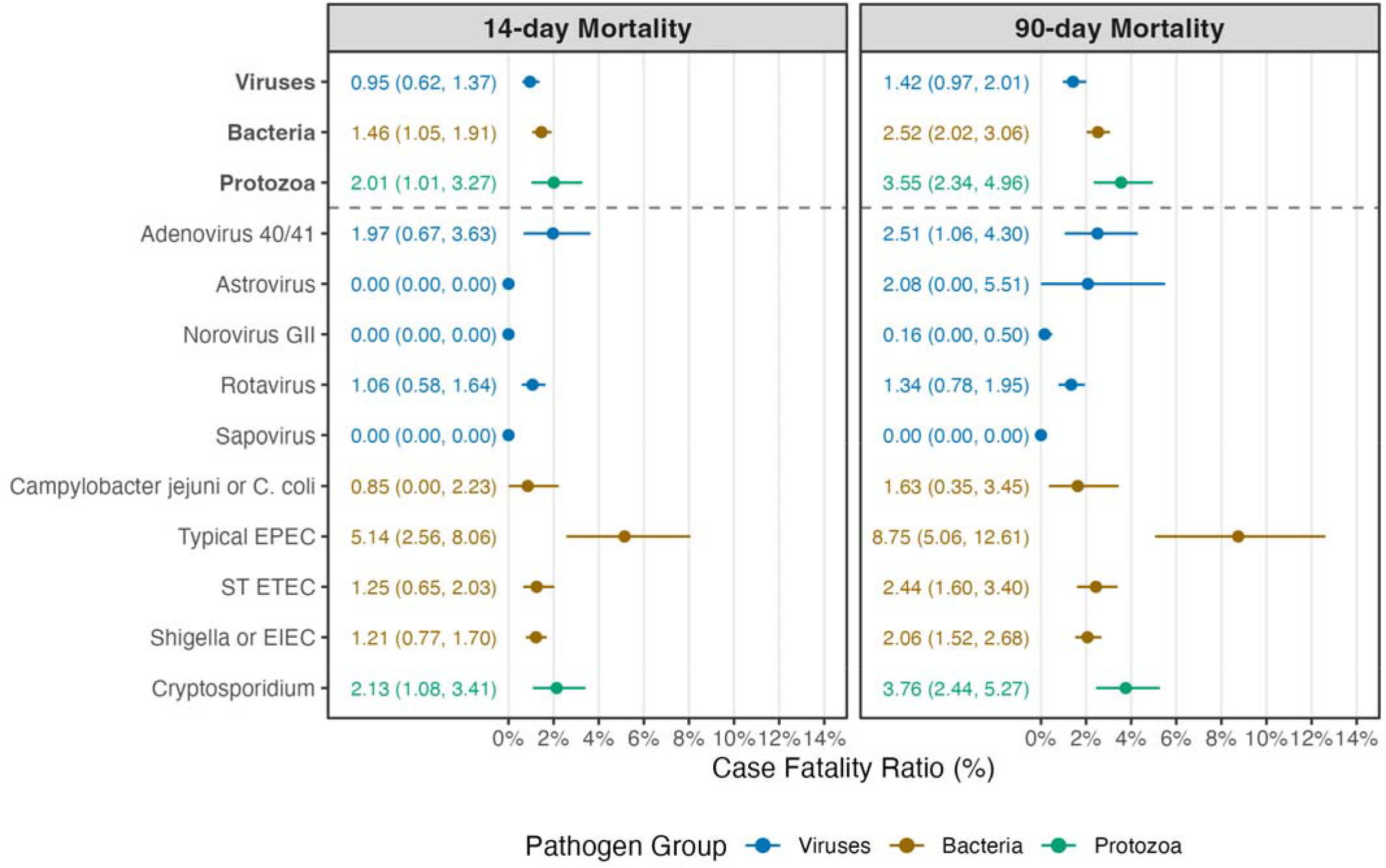
Average 14- and 90-day CFR across studies for individual etiologies and groups of etiologies. CFRs for bacteria also include *Salmonella, V. cholerae*, and *Aeromonas*, and CFRs for protozoa also include *Isospora, E. histolytica*, and *Cyclospora*. Studies include GEMS, VIDA, ABCD, and EFGH.

## 4 Discussion

Our harmonized analysis of five major multisite studies of diarrhea conducted between 2007 and 2024 (GEMS, MAL-ED, VIDA, ABCD, and EFGH) provides an overview of temporal and geographic trends in etiology-specific MSD among children in low-resource settings. By aligning case definitions and analytic approaches – including healthcare-seeking adjustments and qPCR-based attribution cutoffs – we were able to estimate more comparable incidence, AFs, and CFRs across studies than what has been previously published. We found that MSD incidence is consistently highest among infants 6–11 months old, with the leading etiology in that age group being rotavirus or *Shigella*, depending on whether it was pre-or post-introduction of rotavirus vaccination into the national immunization program. For children 12-35 months of age, *Shigella* was consistently the leading etiology across country sites and studies. Diarrhea mortality has declined over time, but there appears to be important heterogeneity between etiology-specific CFRs. Our findings are generally consistent with prior analyses of these studies.^8–10,12,15^

Prior to rotavirus vaccine introduction, we found that rotavirus was the leading etiology among children 6-11 months old causing about a quarter of all MSD cases, with the highest rotavirus incidence also in that age group. Our results suggest that, following national rotavirus vaccine introduction, there have been declines in rotavirus MSD. These declines parallel the extensive global evidence showing that rotavirus vaccination has significantly reduced severe rotavirus disease and hospitalizations in both high-income and LMIC settings.^18–20^ However, one site in the studies included (Kenya) did not show clear declines in rotavirus incidence following vaccine introduction when comparing across studies, which may be due to differences between the catchment area populations across studies.^21^ Prior studies have shown high effectiveness of rotavirus vaccination in Kenya.^22,23^

Regardless of rotavirus vaccine availability, we found that *Shigella* was consistently the leading etiology among children 12-35 months of age, causing between a third and half of all MSD in African and Asian sites. Across country sites and studies, *Shigella* incidence peaked at 12-23 months of age and then declined. *Shigella* also replaced rotavirus as the leading etiology for children 6-11 months old in many settings where rotavirus vaccine has been introduced, though there was some heterogeneity by site. This is consistent with findings from the Global Pediatric Diarrhea Surveillance network, which enrolls children <5 years hospitalized with diarrhea in 28 low- and middle-income countries, and has found *Shigella* to be a leading cause of diarrhea hospitalization, second only to rotavirus.^24^

Overall 90-day mortality declined over time, from 2.21% in GEMS^25^ to 0.30% in EFGH,^15^ consistent with observed declines in pediatric diarrhea mortality globally.^26^ The declines across studies may be attributed to improvements in WASH, access to ORS, and rotavirus vaccine availability, but may also be influenced by other factors including 1) Hawthorne effect by repeated use of the same populations across studies, 2) varying protocol driven interventions (e.g., rehydration required in ABCD, antibiotic use in EFGH), and 3) other factors such as varying rotavirus vaccine coverage and socioeconomic status in sites across studies.

Other than GEMS, the data included was too sparse to estimate study-, age-, and etiology-specific CFRs. Therefore, we estimated etiology-specific CFRs across studies and age groups, which obscures potential declines in etiology-specific CFRs over time as well as differences between age groups, but elucidates relative risk of mortality across pathogens. We found that typical EPEC had the highest 90-day CFR at 8.75%. 90-day CFRs for other pathogens ranged from 0% (sapovirus) to 3.76% (*Cryptosporidium*) but precision varied and confidence intervals often overlapped. On average, bacteria and protozoa had higher CFRs than viruses, consistent with previous analyses.^25^

Despite our best efforts to harmonize case definitions and analytic methodology across studies, there are some important limitations to consider when comparing results across studies. First, MAL-ED followed a fundamentally different study design than the other studies, actively surveilling a cohort in the community. On the other hand, GEMS, VIDA, and EFGH only enrolled cases who sought care at healthcare facilities. To minimize this limitation, we adjusted for healthcare seeking in GEMS, VIDA, and EFGH using previously validated methodology that accounts for heterogeneity in care seeking by age, disease severity, and socioeconomic status.^14^ Second, the population catchment area within each country often varied across studies. For example, in Bangladesh, GEMS enrolled cases in Mirzapur, a relatively rural setting, whereas MAL-ED, ABCD, and EFGH enrolled cases in Dhaka, the country’s capital. These populations differ substantially, so the apparent decline in *Shigella* and overall MSD incidence from GEMS to VIDA and EFGH in Bangladesh is likely to be affected by these differences. Third, some unresolvable differences remained in our harmonization of analytic methodology across studies. For example, we were not able to incorporate socioeconomic status when adjusting for care seeking in GEMS, and the HUS survey in VIDA asked participants about diarrhea episodes in the past week vs two weeks in GEMS and EFGH.^16^

In conclusion, despite some heterogeneity across studies and country sites, our results show strong and consistent evidence of the importance of individual etiologies in the burden of diarrhea among children in low-resource settings. Among the youngest children, rotavirus continues to be a leading cause of MSD, particularly in settings where rotavirus vaccination has not yet been introduced or scaled up. For children over 1 year old, *Shigella* is the dominant cause of MSD. Our findings support continued efforts to introduce rotavirus vaccination globally, as well as to develop and introduce a *Shigella* vaccine. Ongoing high-quality, harmonized surveillance will be crucial for monitoring future shifts in etiology, guiding vaccine policy, and accelerating progress in reducing global diarrheal morbidity and mortality.

## Data Availability

GEMS data are available online through ClinEpiDB, including GEMS1 Case Control data (https://clinepidb.org/ce/app/workspace/analyses/DS_841a9f5259/new) and GEMS1 HUAS/HUAS Lite data (https://clinepidb.org/ce/app/workspace/analyses/DS_221d2bcac4/new). MAL-ED data are also available online through ClinEpiDB (https://clinepidb.org/ce/app/workspace/analyses/DS_5c41b87221/new). VIDA data are available through ClinEpiDB by request to the study team. ABCD data are available by request to the study team. EFGH data can be found in Vilvi (https://doi.org/10.25934/PR00011860).

https://mariagq.shinyapps.io/diar-trends-dashboard/

https://clinepidb.org/ce/app/workspace/analyses/DS_841a9f5259/new

https://clinepidb.org/ce/app/workspace/analyses/DS_221d2bcac4/new

https://clinepidb.org/ce/app/workspace/analyses/DS_5c41b87221/new

https://doi.org/10.25934/PR00011860

## References

1. Kyu HH, Vongpradith A, Dominguez RMV, Ma J, Albertson SB, Novotney A, et al. Global, regional, and national age-sex-specific burden of diarrhoeal diseases, their risk factors, and aetiologies, 1990–2021, for 204 countries and territories: a systematic analysis for the Global Burden of Disease Study 2021. Lancet Infect Dis. 2024 Dec 18;0(0). doi:10.1016/S1473-3099(24)00691-1 PubMed PMID: 39708822.

2. Naghavi M, Ong KL, Aali A, Ababneh HS, Abate YH, Abbafati C, et al. Global burden of 288 causes of death and life expectancy decomposition in 204 countries and territories and 811 subnational locations, 1990–2021: a systematic analysis for the Global Burden of Disease Study 2021. The Lancet. 2024 Apr 3;0(0). doi:10.1016/S0140-6736(24)00367-2 PubMed PMID: 38582094.

3. Kotloff KL, Blackwelder WC, Nasrin D, Nataro JP, Farag TH, van Eijk A, et al. The Global Enteric Multicenter Study (GEMS) of diarrheal disease in infants and young children in developing countries: epidemiologic and clinical methods of the case/control study. Clin Infect Dis Off Publ Infect Dis Soc Am. 2012 Dec;55 Suppl 4(Suppl 4):S232–245. doi:10.1093/cid/cis753 PubMed PMID: 23169936; PubMed Central PMCID: PMC3502307.

4. MAL-ED Network Investigators. The MAL-ED study: a multinational and multidisciplinary approach to understand the relationship between enteric pathogens, malnutrition, gut physiology, physical growth, cognitive development, and immune responses in infants and children up to 2 years of age in resource-poor environments. Clin Infect Dis Off Publ Infect Dis Soc Am. 2014 Nov 1;59 Suppl 4:S193-206. doi:10.1093/cid/ciu653 PubMed PMID: 25305287.

5. Powell H, Liang Y, Neuzil KM, Jamka LP, Nasrin D, Sow SO, et al. A Description of the Statistical Methods for the Vaccine Impact on Diarrhea in Africa (VIDA) Study. Clin Infect Dis. 2023 Apr 1;76(Supplement_1):S5–11. doi:10.1093/cid/ciac968

6. Vannice K, MacLennan CA, Long J, Steele AD. Optimizing Vaccine Trials for Enteric Diseases: The Enterics for Global Health (EFGH) Shigella Surveillance Study. Open Forum Infect Dis. 2024 Mar 1;11(Supplement_1):S1–5. doi:10.1093/ofid/ofad586

7. Blackwelder WC, Biswas K, Wu Y, Kotloff KL, Farag TH, Nasrin D, et al. Statistical Methods in the Global Enteric Multicenter Study (GEMS). Clin Infect Dis Off Publ Infect Dis Soc Am. 2012 Dec 15;55(Suppl 4):S246–53. doi:10.1093/cid/cis788 PubMed PMID: 23169937; PubMed Central PMCID: PMC3502316.

8. Liu J, Platts-Mills JA, Juma J, Kabir F, Nkeze J, Okoi C, et al. Use of quantitative molecular diagnostic methods to identify causes of diarrhoea in children: a reanalysis of the GEMS case-control study. The Lancet. 2016 Sep 24;388(10051):1291–301. doi:10.1016/S0140-6736(16)31529-X

9. Nasrin D, Liang Y, Powell H, Casanova IG, Sow SO, Hossain MJ, et al. Moderate-to-Severe Diarrhea and Stunting Among Children Younger Than 5 Years: Findings From the Vaccine Impact on Diarrhea in Africa (VIDA) Study. Clin Infect Dis. 2023 Apr 1;76(Supplement_1):S41–8. doi:10.1093/cid/ciac945

10. Platts-Mills JA, Liu J, Rogawski ET, Kabir F, Lertsethtakarn P, Siguas M, et al. Use of quantitative molecular diagnostic methods to assess the aetiology, burden, and clinical characteristics of diarrhoea in children in low-resource settings: a reanalysis of the MAL-ED cohort study. Lancet Glob Health. 2018 Dec;6(12):e1309–18. doi:10.1016/S2214-109X(18)30349-8

11. The Antibiotics for Children With Diarrhea (ABCD) Study Group, Ahmed T, Chisti MJ, Rahman MW, Alam T, Ahmed D, et al. Effect of 3 Days of Oral Azithromycin on Young Children With Acute Diarrhea in Low-Resource Settings: A Randomized Clinical Trial. JAMA Netw Open. 2021 Dec 16;4(12):e2136726. doi:10.1001/jamanetworkopen.2021.36726

12. Pavlinac PB, Platts-Mills J, Liu J, Atlas HE, Gratz J, Operario D, et al. Azithromycin for bacterial watery diarrhea: A reanalysis of the AntiBiotics for Children with severe Diarrhea (ABCD) trial incorporating molecular diagnostics. J Infect Dis. 2023 Jul 5;jiad252. doi:10.1093/infdis/jiad252 PubMed PMID: 37405406.

13. Liu J, Garcia Bardales PF, Islam K, Jarju S, Juma J, Mhango C, et al. Shigella Detection and Molecular Serotyping With a Customized TaqMan Array Card in the Enterics for Global Health (EFGH): Shigella Surveillance Study. Open Forum Infect Dis. 2024 Mar 1;11(Supplement_1):S34–40. doi:10.1093/ofid/ofad574

14. Garcia Quesada M, Breskin A, Platts-Mills JA, Benkeser D, Pavlinac PB, Galagan SR, et al. Comparing existing and novel methods for estimating etiology-specific diarrheal disease incidence in hybrid surveillance studies [Internet]. medRxiv; 2025 [cited 2026 Jan 25]. p. 2025.11.11.25339698. Available from: https://www.medrxiv.org/content/10.1101/2025.11.11.25339698v1 doi:10.1101/2025.11.11.25339698

15. Yousafzai MT, Cornick J, Penataro Yori P, Hossain MJ, Keita AM, Atlas HE, et al. The incidence and antimicrobial resistance of Shigella-attributable diarrhoea in young children in low-income and middle-income countries from the multicountry Enterics for Global Health (EFGH) Shigella Surveillance Study: a prospective, facility-based hybrid surveillance study. Lancet Glob Health. 2026 Mar 11. doi:10.1016/S2214-109X(25)00534-0

16. Nasrin D, Wu Y, Blackwelder WC, Farag TH, Saha D, Sow SO, et al. Health care seeking for Childhood Diarrhea in Developing Countries: Evidence from Seven Sites in Africa and Asia. Am J Trop Med Hyg. 2013 Jul 10;89(1_Suppl):3–12. doi:10.4269/ajtmh.12-0749

17. Dodd R, Awuor AO, Garcia Bardales PF, Khanam F, Mategula D, Onwuchekwa U, et al. Population Enumeration and Household Utilization Survey Methods in the Enterics for Global Health (EFGH): Shigella Surveillance Study. Open Forum Infect Dis. 2024 Mar 1;11(Supplement_1):S17–24. doi:10.1093/ofid/ofae018

18. Aliabadi N, Antoni S, Mwenda JM, Weldegebriel G, Biey JNM, Cheikh D, et al. Global impact of rotavirus vaccine introduction on rotavirus hospitalisations among children under 5 years of age, 2008–16: findings from the Global Rotavirus Surveillance Network. Lancet Glob Health. 2019 Jul 1;7(7):e893–903. doi:10.1016/S2214-109X(19)30207-4 PubMed PMID: 31200889.

19. Jonesteller CL, Burnett E, Yen C, Tate JE, Parashar UD. Effectiveness of Rotavirus Vaccination: A Systematic Review of the First Decade of Global Postlicensure Data, 2006– 2016. Clin Infect Dis. 2017 Sep 1;65(5):840–50. doi:10.1093/cid/cix369

20. Troeger C, Khalil IA, Rao PC, Cao S, Blacker BF, Ahmed T, et al. Rotavirus Vaccination and the Global Burden of Rotavirus Diarrhea Among Children Younger Than 5 Years. JAMA Pediatr. 2018 Oct 1;172(10):958–65. doi:10.1001/jamapediatrics.2018.1960

21. Amin AB, Waller LA, Tate JE, Lash TL, Lopman BA. Accounting for local incidence when estimating rotavirus vaccine efficacy among countries: a pooled analysis of monovalent rotavirus vaccine trials. Am J Epidemiol. 2025 Nov 4;194(11):3168–74. doi:10.1093/aje/kwae467

22. Khagayi S, Omore R, Otieno GP, Ogwel B, Ochieng JB, Juma J, et al. Effectiveness of Monovalent Rotavirus Vaccine Against Hospitalization With Acute Rotavirus Gastroenteritis in Kenyan Children. Clin Infect Dis. 2020 May 23;70(11):2298–305. doi:10.1093/cid/ciz664

23. Wandera EA, Kurokawa N, Mutua MM, Muriithi B, Nyangao J, Khamadi SA, et al. Long-term impact of rotavirus vaccination on all-cause and rotavirus-specific gastroenteritis and strain distribution in Central Kenya: An 11-year interrupted time-series analysis. Vaccine. 2024 Sep 17;42(22):126210. doi:10.1016/j.vaccine.2024.126210

24. Cohen AL, Platts-Mills JA, Nakamura T, Operario DJ, Antoni S, Mwenda JM, et al. Aetiology and incidence of diarrhoea requiring hospitalisation in children under 5 years of age in 28 low-income and middle-income countries: findings from the Global Pediatric Diarrhea Surveillance network. BMJ Glob Health. 2022 Sep;7(9):e009548. doi:10.1136/bmjgh-2022-009548 PubMed PMID: 36660904; PubMed Central PMCID: PMC9445824.

25. Levine MM, Nasrin D, Acácio S, Bassat Q, Powell H, Tennant SM, et al. Diarrhoeal disease and subsequent risk of death in infants and children residing in low-income and middle-income countries: analysis of the GEMS case-control study and 12-month GEMS-1A follow-on study. Lancet Glob Health. 2020 Feb 1;8(2):e204–14. doi:10.1016/S2214-109X(19)30541-8 PubMed PMID: 31864916.

26. Liu L, Oza S, Hogan D, Perin J, Rudan I, Lawn JE, et al. Global, regional, and national causes of child mortality in 2000–13, with projections to inform post-2015 priorities: an updated systematic analysis. The Lancet. 2015 Jan 31;385(9966):430–40. doi:10.1016/S0140-6736(14)61698-6

27. International Vaccine Access Center (IVAC), Johns Hopkins Bloomberg School of Public Health. VIEW-hub [Internet]. [cited 2026 Jan 15]. Rotavirus Vaccine. Available from: https://view-hub.org/vaccine/rota

