## Supplementary Tables and Figures for "Geographic and temporal trends in etiology-specific diarrhea burden among children in low-resource settings"

**Supplementary tables & figures for: "Geographic and temporal trends in etiology-specific diarrhea burden among children in LMICs"**

Supplementary Table 1. Incidence per 100 person-years of moderate-to-severe diarrhea (MSD) by age group, study, and country site.

| ***Study*** | ***Region*** | ***Country*** | ***0-5 months*** | ***6-11 months*** | ***12-23 months*** | ***24-35 months*** | ***36-59 months*** | ***Rotavirus Vaccine**** |
|---|---|---|---|---|---|---|---|---|
| GEMS | Africa | Kenya | 44.69 (38.31, 52.19) | 70.88 (62.89, 80.06) | 23.79 (21.37, 26.65) | 12.08 (10.49, 13.96) | 5.00 (4.22, 5.93) | No |
| | | Mali | 42.77 (29.54, 73.22) | 119.12 (92.85, 186.47) | 64.86 (50.98, 103.94) | 29.83 (21.31, 53.28) | 12.50 (7.25, 29.33) | No |
| | | Mozambique | 13.34 (10.15, 18.76) | 21.46 (17.34, 28.42) | 13.37 (10.63, 17.44) | 6.10 (4.07, 9.71) | 1.50 (0.82, 2.93) | No |
| | | The Gambia | 5.55 (3.93, 7.59) | 25.73 (22.28, 30.67) | 17.81 (15.90, 20.95) | 5.58 (4.53, 6.91) | 0.95 (0.65, 1.34) | No |
| | Asia | Bangladesh | 18.01 (11.82, 34.29) | 44.27 (32.07, 76.66) | 21.91 (16.70, 36.02) | 10.28 (7.57, 18.15) | 3.55 (2.15, 8.24) | No |
| | | India | 49.48 (38.67, 68.40) | 102.00 (83.34, 131.81) | 65.08 (52.28, 84.21) | 21.10 (16.08, 28.47) | 9.37 (6.61, 14.74) | No |
| | | Pakistan | 24.43 (20.07, 30.18) | 33.59 (28.25, 41.47) | 16.91 (14.23, 20.63) | 6.24 (4.97, 8.06) | 2.26 (1.67, 2.96) | No |
| MAL-ED | Africa | Tanzania | 14.35 (8.65, 23.81) | 14.35 (8.65, 23.81) | 1.44 (0.46, 4.45) | — | — | Partial |
| | | South Africa | 0.84 (0.12, 5.99) | 1.69 (0.42, 6.75) | 1.27 (0.41, 3.92) | — | — | Yes |
| | Asia | Bangladesh | 20.95 (13.80, 31.82) | 51.43 (39.39, 67.15) | 26.19 (20.11, 34.11) | — | — | No |
| | | India | 5.29 (2.37, 11.77) | 16.74 (10.68, 26.24) | 16.30 (11.81, 22.50) | — | — | No |
| | | Nepal | 7.96 (4.14, 15.31) | 32.60 (23.62, 44.99) | 18.50 (13.67, 25.04) | — | — | No |
| | | Pakistan | 108.13 (91.23, 128.16) | 106.50 (89.74, 126.40) | 68.29 (58.71, 79.44) | — | — | No |
| | Latin America | Brazil | 1.23 (0.17, 8.71) | 0.00 (0.00, Inf) | 0.61 (0.09, 4.30) | — | — | Yes |
| | | Peru | 25.77 (17.42, 38.14) | 75.26 (59.83, 94.66) | 70.62 (59.73, 83.49) | — | — | Yes |
| VIDA | Africa | Kenya | 16.47 (13.49, 20.12) | 26.04 (22.60, 30.77) | 19.87 (17.56, 22.86) | 8.85 (7.45, 10.38) | 3.36 (2.75, 4.29) | Yes |
| | | Mali | 21.84 (11.41, 61.44) | 77.73 (48.18, 201.53) | 69.44 (45.01, 182.64) | 57.52 (31.65, 182.04) | 27.16 (9.93, 164.76) | Yes |
| | | The Gambia | 10.44 (7.55, 15.19) | 68.54 (55.80, 87.30) | 51.21 (41.70, 65.00) | 14.15 (11.49, 17.90) | 2.55 (1.80, 3.68) | Yes |
| EFGH | Africa | Kenya | — | 86.17 (59.17, 141.68) | 46.61 (33.18, 74.18) | 24.89 (16.99, 43.94) | — | Yes |
| | | Mali | — | 21.94 (13.10, 44.24) | 10.46 (6.95, 20.03) | 3.02 (1.57, 6.75) | — | Yes |
| | | The Gambia | — | 20.46 (13.37, 34.00) | 23.21 (15.94, 36.35) | 8.49 (5.61, 14.38) | — | Yes |
| | | Malawi | — | 5.36 (3.57, 8.50) | 4.44 (3.18, 6.90) | 2.85 (1.84, 4.97) | — | Yes |
| | Asia | Bangladesh | — | 17.36 (13.68, 24.85) | 9.64 (7.43, 14.78) | 3.76 (2.39, 6.87) | — | No |
| | | Pakistan | — | 14.66 (10.96, 20.19) | 12.91 (9.94, 17.74) | 4.54 (3.23, 6.58) | — | Yes |
| | Latin America | Peru | — | 145.69 (96.01, 335.85) | 132.89 (98.26, 276.21) | 46.80 (29.50, 121.90) | — | Yes |

*Rotavirus vaccine introduction status into national immunization program. No = vaccine not introduced at the time of the study. Partial = vaccine introduced during the study. Yes = vaccine introduced prior to the start of the study.

Supplementary Table 2. Average attributable fractions (AFs) across studies and countries pre- and post-rotavirus vaccine introduction into the national immunization program, by age group.

| ***Region*** | ***Rotavirus Vaccine**** | ***Pathogen Group*** | ***Pathogen*** | ***6-11 months*** | ***12-23 months*** | ***24-35 months*** |
|---|---|---|---|---|---|---|
| Africa | No | Viruses | Adenovirus 40/41 | 4.63 (3.46, 6.10) | 3.97 (2.81, 5.42) | 2.29 (0.92, 3.90) |
| | | Viruses | Astrovirus | 3.33 (1.32, 6.52) | 1.37 (0.73, 2.10) | 1.27 (0.46, 2.45) |
| | | Viruses | Norovirus GII | 2.79 (1.76, 4.12) | 0.57 (0.23, 0.98) | 0.33 (0.00, 0.89) |
| | | Viruses | Rotavirus | 22.40 (17.97, 27.17) | 17.17 (9.07, 30.87) | 5.46 (3.61, 7.57) |
| | | Viruses | Sapovirus | 1.24 (0.60, 1.97) | 1.33 (0.78, 1.99) | 1.49 (0.54, 2.70) |
| | | Bacteria | *Campylobacter jejuni or C. coli* | 2.82 (1.71, 4.03) | 0.98 (0.47, 1.68) | 0.61 (0.00, 1.57) |
| | | Bacteria | *Shigella or EIEC* | 16.36 (11.63, 21.87) | 36.65 (27.79, 50.71) | 37.92 (32.96, 43.21) |
| | | Bacteria | Typical EPEC | 4.03 (1.99, 6.80) | 1.45 (0.76, 2.41) | 0.61 (0.00, 1.68) |
| | | Bacteria | ST ETEC | 14.54 (9.68, 19.69) | 9.74 (7.81, 12.47) | 10.19 (7.16, 13.06) |
| | | Protozoa | *Cryptosporidium* | 14.52 (11.18, 18.47) | 7.27 (5.71, 9.30) | 2.36 (1.00, 3.99) |
| | Yes | Viruses | Adenovirus 40/41 | 3.30 (2.50, 4.29) | 3.71 (2.54, 5.25) | 1.96 (1.07, 3.01) |
| | | Viruses | Astrovirus | 1.63 (1.08, 2.28) | 8.12 (1.49, 14.83) | 1.04 (0.45, 1.92) |
| | | Viruses | Norovirus GII | 4.22 (3.12, 5.52) | 1.59 (1.07, 2.45) | 0.17 (0.04, 0.37) |
| | | Viruses | Rotavirus | 9.74 (8.07, 12.11) | 8.28 (6.81, 10.39) | 8.63 (5.84, 12.08) |
| | | Viruses | Sapovirus | 1.82 (1.06, 2.69) | 2.08 (1.51, 2.90) | 0.96 (0.50, 1.52) |
| | | Bacteria | *Campylobacter jejuni or C. coli* | 9.58 (2.77, 16.53) | 1.17 (0.55, 1.95) | 0.98 (0.33, 1.90) |
| | | Bacteria | *Shigella or EIEC* | 16.45 (8.89, 23.98) | 29.70 (27.08, 33.90) | 34.95 (30.75, 39.96) |
| | | Bacteria | Typical EPEC | 3.64 (2.49, 5.13) | 1.20 (0.68, 1.90) | 0.44 (0.06, 1.16) |
| | | Bacteria | ST ETEC | 9.61 (7.75, 12.10) | 11.03 (9.37, 13.36) | 8.64 (6.75, 10.62) |
| | | Protozoa | *Cryptosporidium* | 11.78 (9.81, 14.27) | 6.85 (5.56, 8.41) | 3.96 (2.84, 5.34) |
| Asia | No | Viruses | Adenovirus 40/41 | 4.67 (3.38, 6.03) | 4.78 (3.13, 6.74) | 3.18 (1.77, 4.81) |
| | | Viruses | Astrovirus | 4.81 (3.26, 6.72) | 2.82 (1.72, 4.21) | 1.05 (0.32, 1.88) |
| | | Viruses | Norovirus GII | 3.05 (1.77, 4.55) | 1.87 (1.00, 3.09) | 0.39 (0.00, 1.25) |
| | | Viruses | Rotavirus | 28.35 (24.26, 32.80) | 18.54 (15.74, 21.49) | 9.14 (6.09, 12.67) |
| | | Viruses | Sapovirus | 4.74 (2.99, 6.86) | 4.05 (2.33, 5.91) | 0.55 (0.11, 1.24) |
| | | Bacteria | *Shigella or EIEC* | 16.24 (12.78, 19.87) | 43.62 (39.62, 47.32) | 56.45 (50.41, 62.20) |
| | | Bacteria | *Campylobacter jejuni or C. coli* | 3.60 (1.98, 5.75) | 1.46 (0.78, 2.43) | 0.50 (0.00, 1.19) |
| | | Bacteria | Typical EPEC | 0.99 (0.52, 1.53) | 0.30 (0.07, 0.60) | 0.20 (0.00, 0.65) |
| | | Bacteria | ST ETEC | 7.88 (5.65, 10.61) | 10.03 (7.91, 12.64) | 6.77 (4.35, 10.56) |
| | | Protozoa | *Cryptosporidium* | 4.33 (2.69, 6.30) | 4.66 (3.28, 6.40) | 3.21 (1.56, 5.87) |
| | Yes | Viruses | Adenovirus 40/41 | 6.17 (2.15, 11.43) | 3.13 (0.39, 7.41) | 1.04 (0.00, 3.60) |
| | | Viruses | Astrovirus | 1.89 (0.00, 5.14) | 1.23 (0.00, 3.10) | 4.49 (0.00, 14.47) |
| | | Viruses | Norovirus GII | 4.56 (1.04, 9.29) | 1.95 (0.00, 6.69) | 1.85 (0.00, 6.05) |
| | | Viruses | Rotavirus | 11.05 (5.50, 17.89) | 5.90 (2.73, 10.04) | 2.31 (0.00, 6.00) |
| | | Viruses | Sapovirus | 3.43 (0.00, 9.51) | 2.32 (0.00, 6.35) | 0.00 (0.00, 0.00) |
| | | Bacteria | *Campylobacter jejuni or C. coli* | 2.53 (0.28, 5.83) | 3.12 (0.54, 6.68) | 2.25 (0.00, 7.50) |
| | | Bacteria | *Shigella or EIEC* | 23.22 (15.84, 31.21) | 37.46 (29.49, 45.80) | 34.49 (21.46, 48.62) |
| | | Bacteria | Typical EPEC | 3.53 (0.53, 7.95) | 1.37 (0.00, 3.58) | 2.25 (0.00, 7.50) |
| | | Bacteria | ST ETEC | 9.65 (4.25, 16.23) | 13.17 (7.95, 19.85) | 19.49 (9.26, 31.30) |
| | | Protozoa | *Cryptosporidium* | 4.67 (1.67, 8.54) | 5.28 (1.47, 10.75) | 0.00 (0.00, 0.00) |

| | | | | | | |
|---|---|---|---|---|---|---|
| Latin America | Yes | Viruses | Adenovirus 40/41 | 4.94 (2.01, 8.45) | 3.45 (2.06, 5.11) | 3.79 (0.64, 8.23) |
| | | Viruses | Astrovirus | 2.14 (0.00, 4.93) | 2.58 (1.18, 4.08) | 1.46 (0.00, 5.11) |
| | | Viruses | Norovirus GII | 4.64 (2.13, 8.12) | 1.21 (0.46, 2.37) | 0.00 (0.00, 0.00) |
| | | Viruses | Rotavirus | 2.85 (0.74, 5.61) | 3.08 (1.76, 4.85) | 3.99 (1.67, 9.29) |
| | | Viruses | Sapovirus | 1.58 (0.00, 3.98) | 3.10 (1.62, 4.70) | 0.48 (0.00, 2.97) |
| | | Bacteria | *Campylobacter jejuni or C. coli* | 4.79 (1.82, 8.54) | 0.44 (0.00, 1.18) | 0.00 (0.00, 0.00) |
| | | Bacteria | *Shigella or EIEC* | 10.83 (7.36, 14.45) | 17.42 (14.34, 20.60) | 22.63 (15.07, 31.98) |
| | | Bacteria | Typical EPEC | 0.08 (0.00, 0.46) | 0.00 (0.00, 0.00) | 0.00 (0.00, 0.00) |
| | | Bacteria | ST ETEC | 2.46 (0.27, 5.20) | 3.77 (2.18, 5.69) | 2.43 (0.00, 6.11) |
| | | Protozoa | *Cryptosporidium* | 3.16 (0.85, 6.31) | 3.24 (1.74, 4.99) | 0.68 (0.00, 2.37) |

*Rotavirus vaccine introduction status into national immunization program. No = vaccine had not yet been introduced in study-countries included. Yes = vaccine was introduced in study-countries included. Rotavirus vaccine was introduced during MAL-ED in Tanzania; this study-site was grouped with pre-rotavirus vaccine sites.

Supplementary Table 3. Top ranked pathogen and attributable fraction for each study site and age group, by rotavirus vaccine introduction status.

| ***Rotavirus vaccine introduction status*** | ***Study*** | ***Country*** | ***6-11 months*** | ***12-23 months*** | ***24-35 months*** |
|---|---|---|---|---|---|
| Pre-rotavirus vaccine introduction | GEMS | Bangladesh | Rotavirus (27.26%; 95% CI: 19.06 - 37.33) | *Shigella or EIEC* (64.36%; 95% CI: 54.68 - 73.39) | *Shigella or EIEC* (82.24%; 95% CI: 71.26 - 90.82) |
| | GEMS | India | Rotavirus (34.31%; 95% CI: 26.83 - 41.80) | Rotavirus (28.39%; 95% CI: 22.79 - 34.44) | *Shigella or EIEC* (45.76%; 95% CI: 37.64 - 53.52) |
| | GEMS | Kenya | Rotavirus (19.27%; 95% CI: 13.50 - 25.42) | *Shigella or EIEC* (23.01%; 95% CI: 17.94 - 28.33) | *Shigella or EIEC* (24.55%; 95% CI: 17.97 - 31.65) |
| | GEMS | Mali | *Cryptosporidium* (21.60%; 95% CI: 13.85 - 31.70) | *Shigella or EIEC* (43.79%; 95% CI: 33.97 - 55.51) | *Shigella or EIEC* (30.86%; 95% CI: 22.33 - 41.03) |
| | GEMS | Mozambique | Rotavirus (35.72%; 95% CI: 27.03 - 45.16) | *Shigella or EIEC* (40.71%; 95% CI: 31.07 - 51.05) | *Shigella or EIEC* (53.53%; 95% CI: 39.40 - 68.44) |
| | GEMS | Pakistan | Rotavirus (29.16%; 95% CI: 22.72 - 35.99) | *Shigella or EIEC* (43.58%; 95% CI: 37.56 - 49.92) | *Shigella or EIEC* (42.21%; 95% CI: 33.08 - 51.59) |
| | GEMS | The Gambia | Rotavirus (25.12%; 95% CI: 18.46 - 31.94) | *Shigella or EIEC* (42.40%; 95% CI: 35.50 - 49.27) | *Shigella or EIEC* (42.72%; 95% CI: 33.17 - 52.07) |
| | MAL-ED | Bangladesh | Rotavirus (38.46%; 95% CI: 26.33 - 52.22) | *Shigella or EIEC* (50.91%; 95% CI: 37.94 - 63.76) | -- |
| | MAL-ED | India | *Shigella or EIEC* (29.41%; 95% CI: 12.80 - 54.19) | *Shigella or EIEC* (66.67%; 95% CI: 49.23 - 80.49) | -- |
| | MAL-ED | Nepal | Rotavirus (24.32%; 95% CI: 13.17 - 40.52) | *Shigella or EIEC* (40.48%; 95% CI: 26.86 - 55.74) | -- |
| | MAL-ED | Pakistan | Sapovirus (12.00%; 95% CI: 7.37 - 18.95) | *Shigella or EIEC* (13.33%; 95% CI: 8.94 - 19.42) | -- |
| | MAL-ED | Tanzania* | ST ETEC (33.33%; 95% CI: 14.60 - 59.40) | Rotavirus (33.33%; 95% CI: 4.34 - 84.65) | -- |
| | EFGH | Bangladesh | Rotavirus (48.45%; 95% CI: 38.67 - 58.54) | *Shigella or EIEC* (43.35%; 95% CI: 32.69 - 53.30) | *Shigella or EIEC* (55.56%; 95% CI: 37.75 - 72.90) |
| Post-rotavirus vaccine introduction | MAL-ED | Brazil | [no MSD cases detected] | Astrovirus (0.00%; 95% CI: 0.00 - 100.00) | -- |
| | MAL-ED | Peru | *Campylobacter jejuni or C. coli* (8.57%; 95% CI: 3.90 - 17.79) | *Shigella or EIEC* (33.83%; 95% CI: 26.31 - 42.28) | -- |
| | MAL-ED | South Africa | *Campylobacter jejuni or C. coli* (50.00%; 95% CI: 5.89 - 94.11) | Astrovirus (50.00%; 95% CI: 5.89 - 94.11) | -- |
| | VIDA | Kenya | ST ETEC (11.12%; 95% CI: 7.99 - 14.49) | *Shigella or EIEC* (29.27%; 95% CI: 25.48 - 33.57) | *Shigella or EIEC* (26.35%; 95% CI: 20.75 - 32.31) |
| | VIDA | Mali | *Cryptosporidium* (18.22%; 95% CI: 13.88 - 23.06) | *Shigella or EIEC* (26.91%; 95% CI: 22.59 - 31.19) | *Shigella or EIEC* (25.15%; 95% CI: 19.09 - 31.64) |
| | VIDA | The Gambia | *Shigella or EIEC* (20.72%; 95% CI: 15.25 - 26.41) | *Shigella or EIEC* (48.09%; 95% CI: 41.35 - 54.25) | *Shigella or EIEC* (43.26%; 95% CI: 37.29 - 49.12) |
| | EFGH | Kenya | Rotavirus (11.13%; 95% CI: 7.93 - 14.79) | *Shigella or EIEC* (20.89%; 95% CI: 16.01 - 26.45) | *Shigella or EIEC* (24.98%; 95% CI: 16.95 - 33.32) |
| | EFGH | Malawi | Rotavirus (12.91%; 95% CI: 4.67 - 24.20) | *Shigella or EIEC* (16.93%; 95% CI: 8.20 - 26.23) | *Shigella or EIEC* (24.62%; 95% CI: 12.14 - 39.08) |
| | EFGH | Mali | *Cryptosporidium* (23.26%; 95% CI: 14.14 - 33.04) | *Shigella or EIEC* (43.54%; 95% CI: 30.29 - 55.72) | *Shigella or EIEC* (27.36%; 95% CI: 6.49 - 52.78) |
| | EFGH | Pakistan | *Shigella or EIEC* (23.22%; 95% CI: 15.84 - 31.21) | *Shigella or EIEC* (37.46%; 95% CI: 29.49 - 45.80) | *Shigella or EIEC* (34.49%; 95% CI: 21.46 - 48.62) |
| | EFGH | Peru | *Shigella or EIEC* (15.94%; 95% CI: 11.23 - 21.00) | *Shigella or EIEC* (18.42%; 95% CI: 14.01 - 23.46) | *Shigella or EIEC* (22.63%; 95% CI: 15.07 - 31.98) |
| | EFGH | The Gambia | *Shigella or EIEC* (17.38%; 95% CI: 10.00 - 25.91) | *Shigella or EIEC* (51.93%; 95% CI: 43.82 - 60.80) | *Shigella or EIEC* (72.95%; 95% CI: 58.93 - 86.15) |

* Rotavirus vaccine was introduced during MAL-ED in Tanzania.

Supplementary Table 4. Average case fatality ratios (CFR) for pathogen classes and individual pathogens across studies.

| ***Pathogen Group*** | ***Pathogen*** | ***14-day CFR % (95% CI)*** | ***90-day CFR % (95% CI)*** | ***90-day Hospitalized CFR % (95% CI)*** |
|---|---|---|---|---|
| Viruses | All viruses | 0.95 (0.62, 1.37) | 1.42 (0.97, 2.01) | 3.78 (2.05, 6.10) |
| Bacteria | All bacteria | 1.46 (1.05, 1.91) | 2.52 (2.02, 3.06) | 10.06 (7.64, 12.72) |
| Protozoa | All protozoa | 2.01 (1.01, 3.27) | 3.55 (2.34, 4.96) | 12.95 (7.38, 18.20) |
| Viruses | Adenovirus 40/41 | 1.97 (0.67, 3.63) | 2.51 (1.06, 4.30) | 11.03 (4.35, 19.26) |
| Viruses | Astrovirus | 0.00 (0.00, 0.00) | 2.08 (0.00, 5.51) | 15.08 (0.00, 39.78) |
| Viruses | Norovirus GII | 0.00 (0.00, 0.00) | 0.16 (0.00, 0.50) | 0.00 (0.00, 0.00) |
| Viruses | Rotavirus | 1.06 (0.58, 1.64) | 1.34 (0.78, 1.95) | 2.24 (0.91, 3.76) |
| Viruses | Sapovirus | 0.00 (0.00, 0.00) | 0.00 (0.00, 0.00) | 0.00 (0.00, 0.00) |
| Bacteria | *Campylobacter jejuni or C. coli* | 0.85 (0.00, 2.23) | 1.63 (0.35, 3.45) | 12.18 (2.38, 25.00) |
| Bacteria | ST ETEC | 1.25 (0.65, 2.03) | 2.44 (1.60, 3.40) | 7.59 (3.56, 11.99) |
| Bacteria | *Shigella or EIEC* | 1.21 (0.77, 1.70) | 2.06 (1.52, 2.68) | 9.57 (6.47, 12.95) |
| Bacteria | Typical EPEC | 5.14 (2.56, 8.06) | 8.75 (5.06, 12.61) | 27.85 (15.73, 41.62) |
| Protozoa | *Cryptosporidium* | 2.13 (1.08, 3.41) | 3.76 (2.44, 5.27) | 13.09 (7.85, 19.20) |

CFRs for bacteria also include *Salmonella*, *V. cholerae*, and *Aeromonas*, and CFRs for protozoa also include *Isospora*, *E. histolytica*, and *Cyclospora*. Studies include GEMS, VIDA, ABCD, and EFGH.

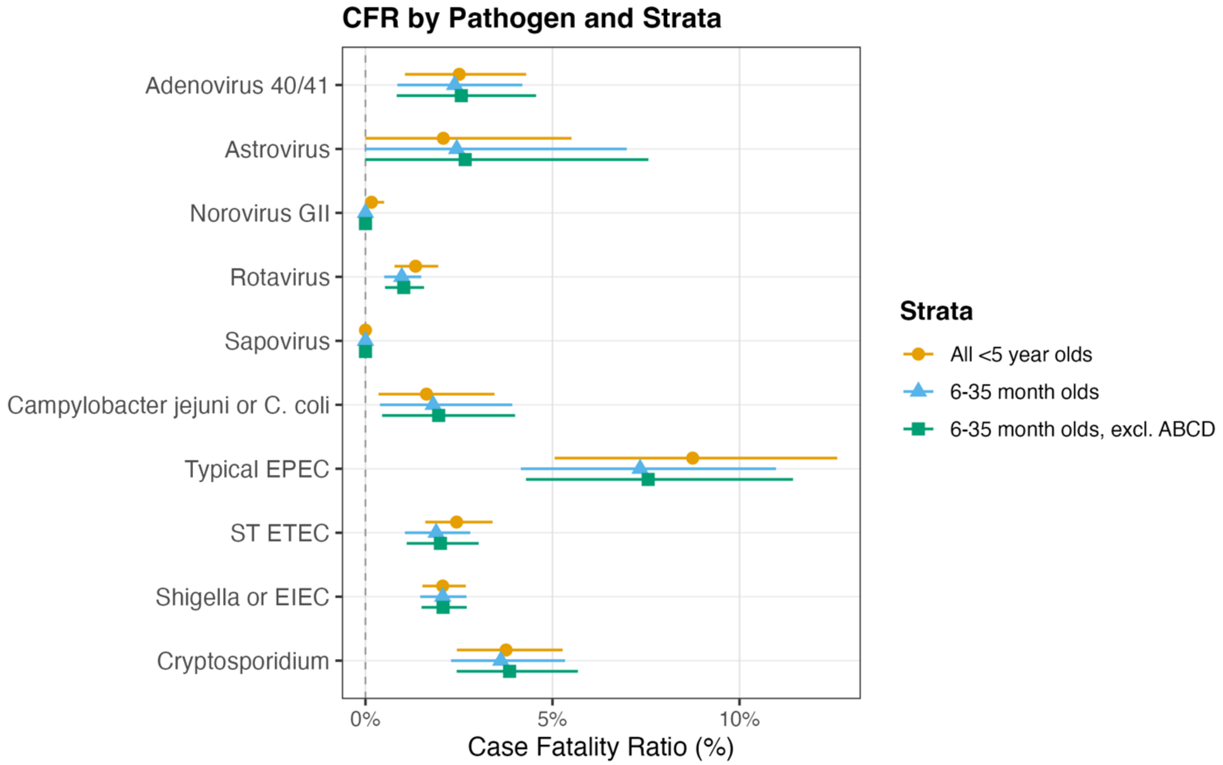

Supplementary Figure 1. Sensitivity analysis of the 90-day CFRs, comparing including the full age range (<5 years), restricting to the age range in EFGH (6-35 months), and excluding ABCD given differing eligibility criteria for cases.
